# Exploring the Molecular Pathways of Intracranial Aneurysm Formation in Autosomal Dominant Polycystic Kidney Disease Using Proteomic Analysis

**DOI:** 10.1101/2024.11.07.24316796

**Authors:** Jin-Myung Kim, Hee-Sung Ahn, Mi Joung Kim, Hye Eun Kwon, Youngmin Ko, Joo Hee Jung, Hyunwook Kwon, Young Hoon Kim, Jiyoung Yu, Kyunggon Kim, Sung Shin

**Affiliations:** Division of Kidney and Pancreas Transplantation, Department of Surgery, Asan Medical Center, University of Ulsan College of Medicine, Seoul, Republic of Korea; AMC sciences, Asan Medical Center, Republic of Korea; Convergence Medicine Research Center, Asan Institute for Life Sciences, Asan Medical Center, Seoul, Republic of Korea; Department of Digital Medicine, BK21 Project, Asan Medical Center, University of Ulsan College of Medicine Seoul, Republic of Korea

## Abstract

**Introduction:** Intracranial aneurysm (IA) frequently coincides with autosomal dominant polycystic kidney disease (ADPKD), exhibiting incidence rates nearly 10 times higher than the general population. However, the exact mechanism of how these two conditions is related remains unclear. This study aims to identify mechanisms behind IA occurrence in ADPKD patients using proteomics and to discover potential protein biomarkers for early diagnosis.

**Method:** Pre-kidney transplantation ADPKD patients underwent cranial CT and/or MR angiography, with findings dictating assignment to either a control group (ADPKD without IA, n=20) or IA group (ADPKD with IA, n=9). During transplantation, bilateral nephrectomy was performed and native renal arteries were sampled for proteomic analysis via a liquid chromatography-tandem mass spectrometry. Differentially expressed proteins were subjected to bioinformatic analysis and a protein-protein interaction network analysis.

**Results:** Eight proteins showed significant variation between IA and control groups, with four proteins upregulated (DIS3, RAB6A, MMS19, EXOC8) and four downregulated (CLUH, SYNC, MEF2D, WDR36) in IA group (Log_2_ fold change (FC) >2 and false discovery rate (FDR] q-value <0.05) compared to the control group. These proteins correlated with pathways implicated in IA development, such as ciliopathy, exocytosis, inflammation, extracellular matrix remodelling, and apoptosis. These proteins were quantitatively validated using Western blot analysis and found to be consistent with proteomic data. Moreover, a connection was observed between protein expression and clinical metrics (bilirubin, prothrombin time, platelet count), indicating their potential as early diagnostic markers.

**Conclusion:** This study is the first to employ renal artery samples to study underlying mechanisms for IA in ADPKD patients by proteomics. We identified and validated novel candidate markers that are either upregulated or downregulated in the IA group compared to the control group. This research’s finding opens new avenues for understanding and diagnosing IA in ADPKD, potentially leading to earlier diagnosis and targeted treatments.

## 1. Introduction

Autosomal dominant polycystic kidney disease (ADPKD) is the most frequent form of hereditary renal disease affecting 1 in 1000 people and characterized by gradual and irreversible decline in renal function while accounting for 10% of cases of the kidney failure^1–3^. Apart from renal manifestations, changes in other organs may be present, including liver cysts and intracranial aneurysm (IA). IAs are a rare vascular manifestation, affecting only 5%–9% of the general population. However, they are substantially more prevalent among ADPKD patients, who experience a three- to five-fold higher incidence rate, with prevalence rates climbing up to 40%^4–6^.

Despite extensive research highlighting a significant association between ADPKD and IAs, the precise mechanisms underlying this relationship are still not fully understood. Although only a limited number of studies have involved large cohorts exceeding 1,000 participants, numerous investigations have focused on identifying risk factors for IA development or establishing guidelines for IA screening in patients with ADPKD^7–9^. One study proposed that vascular remodeling, induced by an escalating inflammatory response, might be a potential mechanism for the development of IAs in ADPKD patients. This hypothesis was supported by the presence of significant inflammatory markers and endothelial dysfunction in affected individuals^10^.

To date, no significant genetic or protein variations have been identified in extensive studies comparing ADPKD patients with IA to those without. This absence of definitive biomarkers has impeded the development of effective screening protocols for detecting IAs in ADPKD patients and has delayed prompt management of the disease before complications arise.

This study aimed to identify protein variations in renal artery samples, using them as surrogate vessels for intracranial vessels, by employing proteomic analysis on two groups of ADPKD patients—those with IA and those without. This approach was predicated on the understanding that ciliopathy^11^, the fundamental pathophysiological mechanism underlying ADPKD, exerts systemic effects on the vascular architecture^12^. The dysfunction of primary cilia, characteristic of ADPKD, is not confined to renal tissues but extends to vascular endothelial and smooth muscle cells, potentially influencing the structural integrity and function of blood vessels throughout the body^13^. The primary objective is to uncover potential underlying mechanisms for IA formation in individuals with ADPKD by identifying differentially expressed proteins through proteomics. The results of this study may provide valuable insights that could aid in monitoring IA development and facilitate early intervention for ADPKD patients.

## 2. METHODS

### 2.1 Patients and sample collection

The present study was conducted following the guidelines of the Declaration of Helsinki and with the approval of the Research Ethics Committee at Asan Medical Center (reference number: 2019-0310). All patients or their legal representatives provided written informed consent before participating in the study. Renal artery samples were collected from a total of 29 individuals from ADPKD patients who underwent kidney transplantation (KT) with concomitant bilateral nephrectomy between December 2018 and June 2022.

At our institution, brain magnetic resonance angiography (MRA) or computed tomography angiography (CTA) are routinely performed on all PCKD patients who are scheduled to undergo KT. Patients who were found to have no aneurysm during the screening were included in the control group. The IA group comprised patients who were found to have aneurysms during screening, including those who received preoperative interventions like coiling or embolization, as well as those who did not undergo these procedures. The detection of IAs involved the identification of a bulging in the intracranial arterial wall measuring at least 1 mm in either CTA or MRA imaging studies^14^. The specimens were rapidly frozen using liquid nitrogen and kept at a temperature of −80°C before analysis.

### 2.2 Pre-operative diagnosis of ADPKD using imaging-based approach

At our center, we do not routinely use genetic testing to identify pathogenic mutations, such as PKD1 and PKD2, in patients suspected of having ADPKD prior to KT, primarily due to the complexity and cost involved. Although genetic testing can help predict the clinical course by identifying the genotype of patients, it is not mandatory to conduct genetic tests before KT solely to confirm ADPKD. Notably, ultrasound has proven to be highly effective in diagnosing ADPKD associated with PKD1 and PKD2 mutations, with an overall sensitivity, specificity, and accuracy of 97%, 100%, and 98%, respectively^15^. Moreover, when ultrasound findings are inconclusive, age-specific MRI criteria offer an additional diagnostic tool^16^. Specifically, the detection of more than 10 renal cysts in patients aged 16 to 40 years yields a 100% positive predictive value and sensitivity for ADPKD diagnosis^17^.

However, to ensure the appropriate selection of candidates for our study, we conducted a post-experiment analysis using serum samples from each patient for genetic testing. This analysis was performed in collaboration with 3billion, Inc. in Seoul, South Korea (https://3billion.io/index), which provided expertise in sample analysis and exome sequencing (detailed methodology available in Supplemental Data 1). Out of the 29 patients, 20 serum samples were submitted for genetic testing, with the most common reason for exclusion being insufficient serum quantity. Among these, 19 samples revealed mutations in either the PKD1 or PKD2 genes. One sample, negative for these mutations, exhibited a single exon deletion, indicating the need for further analysis through single-gene testing. Despite not testing for PKD1 or PKD2 mutations in all samples, the majority of patients diagnosed with ADPCKD through imaging were confirmed by genetic testing, validating our imaging-based diagnostic approach in this study.

### 2.3 Clinical data collection

The medical records of all patients and controls were thoroughly examined in order to collect relevant data pertaining to various factors. Several parameters were examined, including age, gender, BMI, age at brain imaging study, presence of polycystic liver disease (PCLD), history of cerobrovascular accident (CVA), hypertension, diabetes mellitus (DM) and estimated Glomerular Filtration Rate (eGFR). Additionally, we gathered data on the angiographic findings of the aneurysm, including the number of aneurysms, their size (or the size of the largest aneurysm if multiple aneurysms were present), location, and type.

### 2.4 Sample preparation: Protein extraction, enzymatic digestion

The workflow of the present study is illustrated in Figure 1. Firstly, renal arterial tissue samples were resolved in 400 μL of lysed buffer containing 5% SDS, 50 mM triethylammonium bicarbonate (pH 8.5) and 1× Halt™ protease inhibitor cocktail (Thermo Fisher Scientific) and homogenized with passed through a pestle (Kimble™ Kontes™ Pellet Pastle™, Thermo Fisher Scientific) 20 times. Protein extraction was applied by adaptive focused acoustics (AFA) technology (Covaris). The homogenized samples were transferred to the micro TUBE-500 AFA Fiber Screw-Cap (Covaris) and sonicated by S220 Focused-ultrasonicator (Covaris). Samples were subjected to AFA process using 175 peak power, 200 cycles per burst and 10 duty factor for 900 sec at 10 °C. Subsequently, the lysated samples were boiled at 80 °C using a heat block (MaXtable™ H10, Daihan Scientific, Korea) for 10 minutes and centrifuged at 18,000 ×g for 10 minutes at room temperature. We transferred the supernatant fraction to the new lobind tube (Eppendorf). The protein concentration of each supernatant containing extracted proteins was measured with a BCA protein quantification kit (Pierce™ BCA Protein Assay Kit; cat. No.: 23225; Thermo Fisher Scientific, Waltham, MA, USA). A 300 µg aliquot of proteins was dissolved in 50 µL of lysis buffer and added dithiothreitol to a final concentration of 20 mM to the denatured sample, it was incubated at 95 °C for 10 min. The chemically reduced sample was then placed in iodoacetamide at a final concentration of 40 mM and reacted for 30 min at 25 °C in the dark. With a final concentration of 1.2% phosphoric acid, acidified samples were attached to suspension-trapping (S-Trap) mini columns (#CO2-mini-80, ProtiFi, Farmingdale, NY, USA). Following the manufacturer’s protocol, we performed S-Trap proteolysis by adding 12 μg of Lys-C/trypsin mixture (#V5071, Promega, Madison, WI, USA) and incubating at 37 °C for 16 h. The digested peptide mixture was freeze-dried with a cold trap (CentriVap Cold Traps; Labconco, Kansas City, MO, USA) and stored atLJ−LJ80 °C until use.

**Figure 1.**
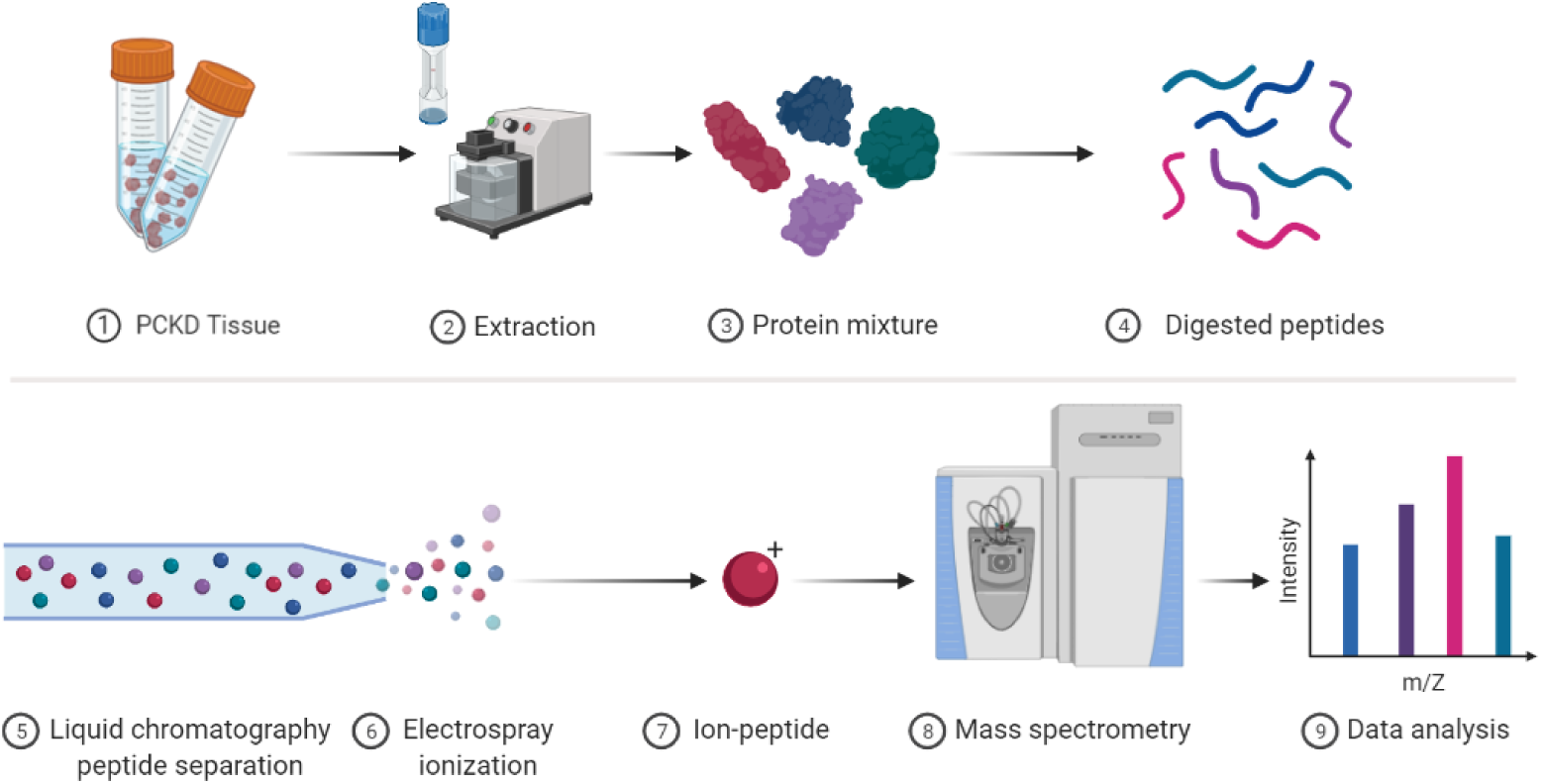
Proteomic analysis workflow of PKCD tissue samples. This schematic diagram outlines the comprehensive steps undertaken in the proteomic analysis of PKCD tissue samples. (A) Sample preparation is initiated with the collection of PKCD tissue (1), followed by protein extraction via mechanical homogenization (2), resulting in a complex protein mixture (3). Subsequent enzymatic digestion breaks down proteins into peptides (4). (B) The analytical phase commences with the separation of peptides using liquid chromatography (5), which are then ionized by electrospray (6). The ionized peptides (7) are introduced into a mass spectrometer (8) that measures their mass-to-charge (m/z) ratios. The resulting data are analyzed to yield a bar graph (9), representing the relative intensities of the detected ions, which facilitates the identification and quantification of peptides present in the sample. Each step is depicted with representative icons and images to illustrate the transition from tissue samples to analyzable molecular data. Figure was created with BioRender.com.

### 2.5 Liquid chromatography-tandem mass spectrometry

Peptide mixtures were separated by using the Dionex UltiMate 3000 RSLC nano system (Thermo Fisher Scientific, Waltham, MA, USA). The mobile phase A was composed of 0.1% FA and 5% DMSO in HPLC-grade water (Avantor, Radnor, PA, USA), while the mobile phase B was made up of 0.1% formic acid (FA), 5% DMSO, and 80% HPLC-grade acetonitrile (Avantor) in HPLC-grade water. The dried sample was resuspended in 0.1% formic acid, and their total peptide concentrations were measured using a UV-Vis spectrophotometer (NanoDrop One, Thermo Fisher Scientific) at a wavelength of 280 nm with the sample type option set to “1 Abs = 1 mg/mL.”. At a concentration of 1μg/μL, 5μL of which was loaded on a C18 Pepmap trap column (20mm×100μm i.d., 5μm, 100Å; Thermo Fisher Scientific) and separated with an Acclaim™ Pepmap 100 C18 column (500mm×75μm i.d., 3μm, 100Å; Thermo Fisher Scientific) over 200min (250nL/min) using a 0–48% acetonitrile gradient in 0.1% formic acid and 5% DMSO for 150min at 50°C. The LC was connected with a Q Exactive HF-X mass spectrometer (Thermo Fisher Scientific) with an EASY-Spray nano-ESI source. In a data-dependent mode, mass spectra were obtained with an automatic switch between a full scan with top 20 data-dependent MS/MS scans. Resolution was set to 60,000 at m/z 200, and target value of 3,000,000 for MS scan type was selected. The ion target value for MS/MS was set at 100,000 with a resolution of 15,000 at m/z 200. The maximum ion injection time was set to 100ms for the full scan and 50ms for MS2 scan. Isolation width was 1.7m/z, and normalized collision energy was set at 27. Dynamic exclusion for measurements of repeated peptides was set for 40s. All mass spectrometry data were measured once per sample and were deposited in the PRIDE archive (www.ebi.ac.uk/pride/archive/projects/PXD043129; Username, Password: 8A6JzKnl)^18^.

### 2.6 Proteomic identification and quantification

The raw files of tandem mass spectrometry (MS/MS) spectra were matched against the UniProtKB/Swiss-Prot human protein sequence database^19^ utilizing SEQUEST HT embedded in Proteome Discoverer (version 2.4; Thermo Fisher Scientific). The search parameters were established at 10 ppm tolerance for precursor ion mass and 0.02 Da for the fragmentation mass. The toleration for trypsin peptides was set at up to two false cleavages, while carbamidomethylation of cysteines was set as a fixed modification, and N-terminal acetylation and methionine oxidation were set as variable modifications. The false discovery rate (FDR) was calculated using the target-decoy search strategy, and the peptides within 1% of the FDR were chosen utilizing the post-processing semi-supervised learning tool Percolator^20^ based on the SEQUEST result. In the global proteome analysis, label-free quantification of proteins was performed using the peak intensity for unique and razor peptides of each protein.

### 2.7 Proteomics data analysis

Raw data were analyzed by Perseus software (version 1.6.15.0)^21^. Log2-transformed raw data were normalized by the width adjustment method. For the comparative statistical analysis, protein selection criteria were based on having a quantified value in 70% of at least on group of two groups: IA group (n=9) and control group (n=20).

### 2.8 Western blot analysis

For the Western blot analysis, proteins were initially lysed using a radioimmunoprecipitation assay (RIPA) buffer supplemented with protease inhibitor cocktails to prevent degradation. We then resolved 10 μg of each protein sample using polyacrylamide gel electrophoresis, which separates proteins based on their molecular weight. Following electrophoresis, the proteins were transferred onto a polyvinylidene difluoride (PVDF) membrane for further analysis. The PVDF membrane was incubated with primary antibodies specific to the target proteins for 1 hour at room temperature to ensure adequate binding. This was followed by a 1-hour incubation with secondary antibodies conjugated to horseradish peroxidase (HRP) at room temperature, which facilitated detection. To visualize the protein bands, we employed an enhanced chemiluminescence (ECL) detection reagent from Bio-Rad Laboratories (Hercules, CA, USA). The resulting chemiluminescent signals were captured using a Bio-Rad Gel Documentation System, providing a clear and accurate representation of the protein expression levels. Finally, we quantified the intensity of the protein bands through densitometry analysis using Image J Software (NIH, Bethesda, MD, USA), allowing us to measure and compare the protein levels accurately.

### 2.9 Statistical analysis

The raw data for the average number of technical replicates for each sample were subjected to a log_2_-transformation and normalized using width adjustment. The different sample groups were compared using Student’s *t*-test and ANOVA tests with Benjamini-Hochberg correction, which was performed using the Perseus software (version 1.6.10.50). The results were then visualized using RStudio (version 1.3.1093), which is a component of R software (version 3.6.0). For all analyses, P<0.05 was considered statistically significant. Several other software packages were also utilized, including PerformanceAnalytics for correlation plotting, ggplot2 for generating boxplots, 2D plots of points and volcano plots. A heatmap was drawn with SRplot (Scientific and Research plot tool)^22^. Statistical significance was defined as a two-tailed test resulting in a p-value less than 0.05, with the inclusion of a false discovery rate (FDR) threshold of less than 0.05.

## 3. Results

### 3.1 Patients characteristics

The study compared various parameters between the control group (n=20) and the IA group (n=9) as shown in Table 1. The mean age was similar in both groups, with the control group at 55.90 ± 6.79 years and the IA group at 57.38 ± 5.68 years (p=0.798). The percentage of females was higher in the IA group (66.7%) compared to the control group (45.0%), though this difference was not statistically significant (p=0.280). Both groups had comparable BMI values, with the control group at 23.66 ± 3.30 kg/m^2^ and the IA group at 24.02 ± 3.52 kg/m^2^ (p=0.554). The age at brain imaging study was also similar, with the control group at 55.53 ± 6.60 years and the IA group at 56.66 ± 6.15 years (p=0.959). The proportion of patients who had dialysis before transplantation was nearly the same in both groups, with 75.0% in the control group and 77.8% in the IA group (p=0.872). However, the duration of hemodialysis was significantly longer in the IA group (21.895 ± 37.91 months) compared to the control group (6.85 ± 11.50 months), with a p-value of 0.001. The presence of PCLD was higher in the IA group (100.0%) than in the control group (75.0%), but this difference was not statistically significant (p=0.099). A history of CVA was present in 22.2% of the IA group and none in the control group (p=0.085). The prevalence of hypertension was slightly higher in the IA group (88.9%) compared to the control group (80.0%), but this difference was not significant (p=0.558). DM was present in 5.0% of the control group and absent in the IA group (p=0.495). Finally, the eGFR was similar in both groups, with the control group at 7.30 ± 2.68 ml/min/1.73m² and the IA group at 6.89 ± 1.53 ml/min/1.73m² (p=0.290). Overall, the significant difference between the two groups was the duration of hemodialysis, which was longer in the IA group.

**Table 1.**
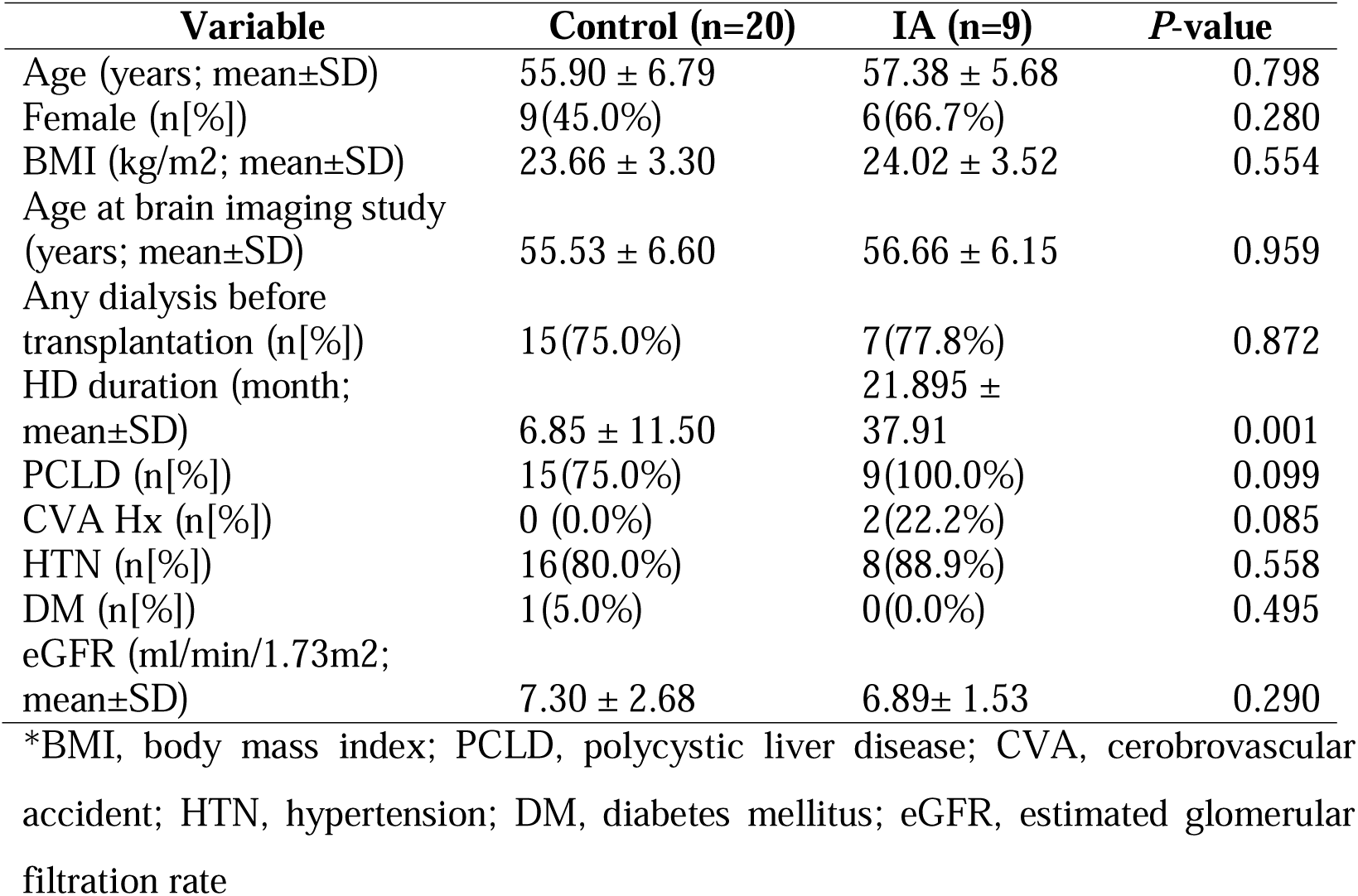
Comparison of demographic and clinical characteristics between control and IA groups.

| Variable | Control (n=20) | IA (n=9) | P-value |
| --- | --- | --- | --- |
| Age (years; mean±SD) | 55.90 ± 6.79 | 57.38 ± 5.68 | 0.798 |
| Female (n[%]) | 9(45.0%) | 6(66.7%) | 0.280 |
| BMI (kg/m<sup>2</sup>; mean±SD) | 23.66 ± 3.30 | 24.02 ± 3.52 | 0.554 |
| Age at brain imaging study (years; mean±SD) | 55.53 ± 6.60 | 56.66 ± 6.15 | 0.959 |
| Any dialysis before transplantation (n[%]) | 15(75.0%) | 7(77.8%) | 0.872 |
| HD duration (month; mean±SD) | 6.85 ± 11.50 | 21.895 ± 37.91 | 0.001 |
| PCLD (n[%]) | 15(75.0%) | 9(100.0%) | 0.099 |
| CVA Hx (n[%]) | 0 (0.0%) | 2(22.2%) | 0.085 |
| HTN (n[%]) | 16(80.0%) | 8(88.9%) | 0.558 |
| DM (n[%]) | 1(5.0%) | 0(0.0%) | 0.495 |
| eGFR (ml/min/1.73m<sup>2</sup>; mean±SD) | 7.30 ± 2.68 | 6.89± 1.53 | 0.290 |

\*BMI, body mass index; PCLD, polycystic liver disease; CVA, cerebrovascular accident; HTN, hypertension; DM, diabetes mellitus; eGFR, estimated glomerular filtration rate

### 3.2 Aneurysm characteristics

The study group consisted of 9 patients with IA, comprising of 3 male and 6 female individuals with an average age of 56.6 years (Table 2). The affected arteries were located as follows: 1 left proximal Posterior Inferior Cerebellar Artery (PICA), 2 right Middle Cerebral Artery (MCA) bifurcations, 1 right Internal Carotid Artery (ICA) ophthalmic branch, 2 Superior Cerebellar Arteries (SCA) (1 on the left, 1 on the right), 1 right distal M1 segment, 1 paraclinoid ICA, and 1 basilar artery top. Each subject had a single IA, with sizes varying from 1mm to 4mm. This data indicates a diverse distribution of aneurysms across different arterial locations within this cohort.

**Table 2.** Aneurysm details in intracranial aneurysm group.

| Subject No. | Age Range | Sex | Intracranial aneurysms |  |  |
| --- | --- | --- | --- | --- | --- |
|  |  |  | Number | Size(mm) | Arteries affected |
| 1 | 60-64 | F | 1 | 2.5 | Left proximal PICA |
| 2 | 60-64 | F | 1 | 2.7 | Right MCA bifurcation |
| 3 | 45-49 | M | 1 | 2 | Right ICA ophthalmic |
| 4 | 50-54 | F | 1 | 4 | Right SCA |
| 5 | 55-59 | F | 1 | 2 | Left SCA |
| 6 | 50-54 | F | 1 | 3 | Right distal M1 |
| 7 | 65-69 | M | 1 | 3 | Right MCA bifurcation |
| 8 | 55-59 | F | 1 | 1 | Paraclinoid ICA |
| 9 | 50-54 | M | 1 | 1 | Basilar artery |

\* PICA, posterior inferior cerebellar artery; MCA, middle cerebral artery; ICA, internal carotid artery; SCA, superior cerebellar artery

### 3.3 Protein identification and differentially expressed proteins across samples

Renal artery samples from each group were pooled for proteomic analysis. We detected and measured 6,223 proteins in at least one sample, with 5,335 of these proteins being quantified in more than 70% of samples within at least one group. Then, protein abundances between IA and control samples were compared using Student’s *t*-tests to identify significant differences, with p-values subsequently adjusted using FDR correction to obtain q-values. A volcano plot, displaying log_2_-fold-changes (IA vs. control) against minus log_10_ q-values, highlighted four proteins that were up-regulated and four proteins that were down-regulated in IA samples compared with the control group (|fold-change| > 1; q-value < 0.05; Figure 2 and Table 3). Among the upregaulted proteins in the IA group, DIS3 (Exosome complex exonuclease RRP44) exhibits the highest upregulation, with a log2 fold-change of 2. RAB6A (Isoform 2 of Ras-related protein Rab-6A) follows closely with a log2 fold-change of 1.97. Other proteins with positive fold-change include MMS19 (MMS19 nucleotide excision repair protein homolog) and EXOC8 (Exocyst complex component 8), with log2 fold-changes of 1.13 and 1.09, respectively.

**Figure 2.**
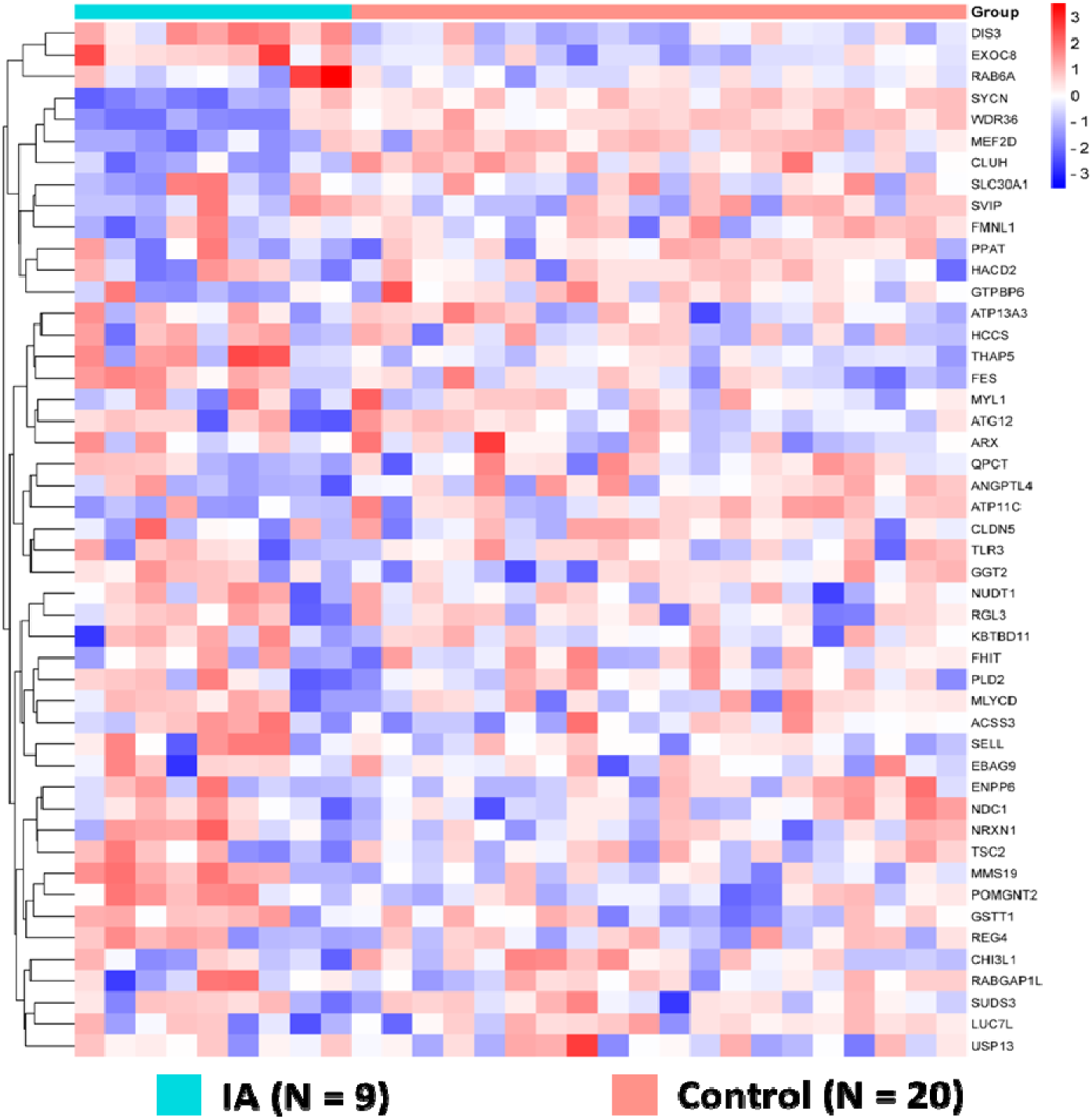
Heatmap visualization of proteomic expression in control and aneurysm groups. The heatmap presents the protein expression profiles across three different groups: control (N = 20), and aneurysm (N = 9). Each row signifies an individual protein, and each column corresponds to a sample within the study groups. The color gradient reflects protein expression levels, with red for upregulation and blue for downregulation. The hierarchical clustering of the samples, based on their similarity or dissimilarity, is depicted by the dendrogram on the left side of the heatmap. As evident from the heatmap, a complex pattern of varying intensities is observed, suggesting the presence of differences or similarities among the samples and features being analyzed

**Table 3.** Comparative analysis of gene expression and protein levels in control vs. intracranial aneurysm group.

| Gene | Protein | FDR *q*-value | Log2 fold-change (A/C) |
| --- | --- | --- | --- |
| DIS3 | Exosome complex exonuclease RRP44 | 0.0451 | 2 |
| RAB6A | Isoform 2 of Ras-related protein Rab-6A | 0.048 | 1.97 |
| MMS19 | MMS19 nucleotide excision repair protein homolog | 0.0107 | 1.13 |
| EXOC8 | Exocyst complex component 8 | 0.044 | 1.09 |
| CLUH | Clustered mitochondria protein homolog | 0.0447 | -1.84 |
| SYCN | Syncollin | 0 | -2.61 |
| MEF2D | Myocyte-specific enhancer factor 2D | 0.011 | -2.93 |
| WDR36 | WD repeat-containing protein 36 | 0 | -3.97 |

Conversely, negative log2 fold-change values indicate proteins that are more highly expressed in the control group, suggesting downregulation in the IA group. CLUH (Clusted mitochondria protein homolog), SYCN (Syncollin) and MEF2D (Myocyte-specific enhancer factor 2D) display significant downregulation with log2 fold-changes of −1.84, −2.61 and −2.93, respectively. The most substantial downregulation is observed in WDR36 (WD repeat-containing protein 36), with a log2 fold-change of −3.97.

The FDR q-value serves as an indicator of statistical significance. SYCN and WDR36, with FDR q-values of 0, demonstrate the highest confidence in differential expression. Other proteins with significant FDR q-values include DIS3 (0.0451), RAB6A (0.048), MMS19 (0.0107), EXOC8 (0.044), and CLUH (0.0447).

In addition, association network in Figure 3 illustrates the protein-protein interaction network for proteins that are either upregulated or downregulated in the IA group relative to the control group, showcasing the interconnected relationships among these differentially expressed proteins. These visual tools effectively depict the proteomic landscape and facilitate the identification of key proteins involved in IA.

**Figure 3.**
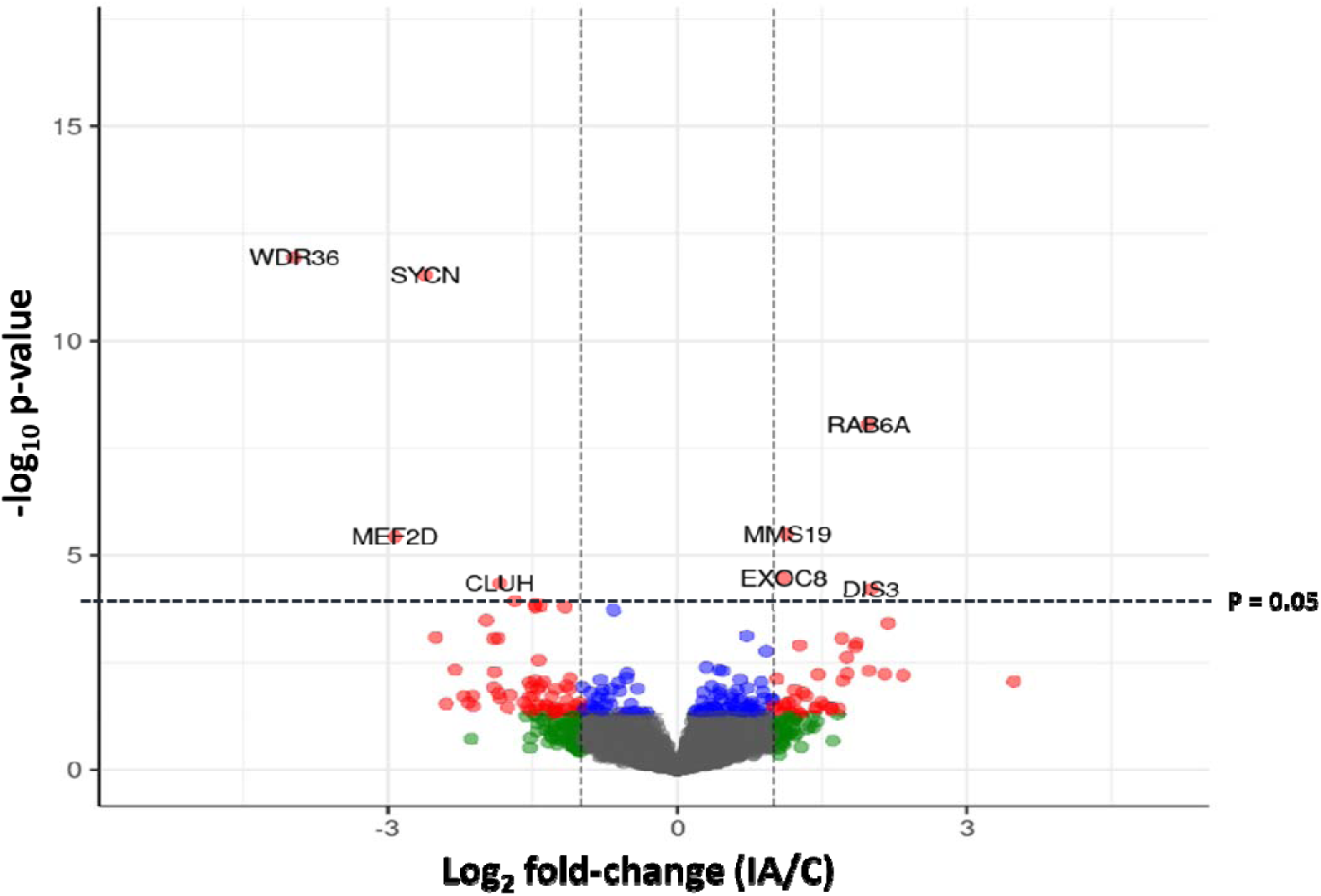
Differential protein expression analysis in aneurysm versus control groups via volcano plot. The volcano plot illustrates the differential expression of proteins between aneurysm (IA) and control (C) groups from proteomic data. Log2 fold-change values (A/C) are plotted on the x-axis, indicating the magnitude of expression changes, while the negative log10 *p*-values are plotted on the y-axis, reflecting statistical significance. The horizontal dashed line represents the significance threshold for the *p*-values, while the vertical dashed lines indicate the fold-change expression cutoffs. Proteins falling outside these thresholds are considered differentially expressed. Statistically significant upregulated proteins in the aneurysm group include MMS19, EXOC8, DIS3 and RAB6a whereas key downregulated proteins comprise WDR36, SYCN, MEF2D, and CLUH.

### 3.6 Heatmap, Volcano Plot and Association Network Analyses

A heatmap clustering analysis was used to demonstrate proteomic diversity between the groups, with distinct expression profiles for the control and IA groups (Figure 2). The volcano plot illustrated the differential expression of proteins, highlighting significant upregulation and downregulation in the IA group compared to controls (Figure 3). These visual tools effectively depict the proteomic landscape and facilitate the identification of key proteins involved in IA. In addition, association network in Figure 4 illustrates the protein-protein interaction network for proteins that are either upregulated or downregulated in the IA group relative to the control group, showcasing the interconnected relationships among these differentially expressed proteins.

**Figure 4.**
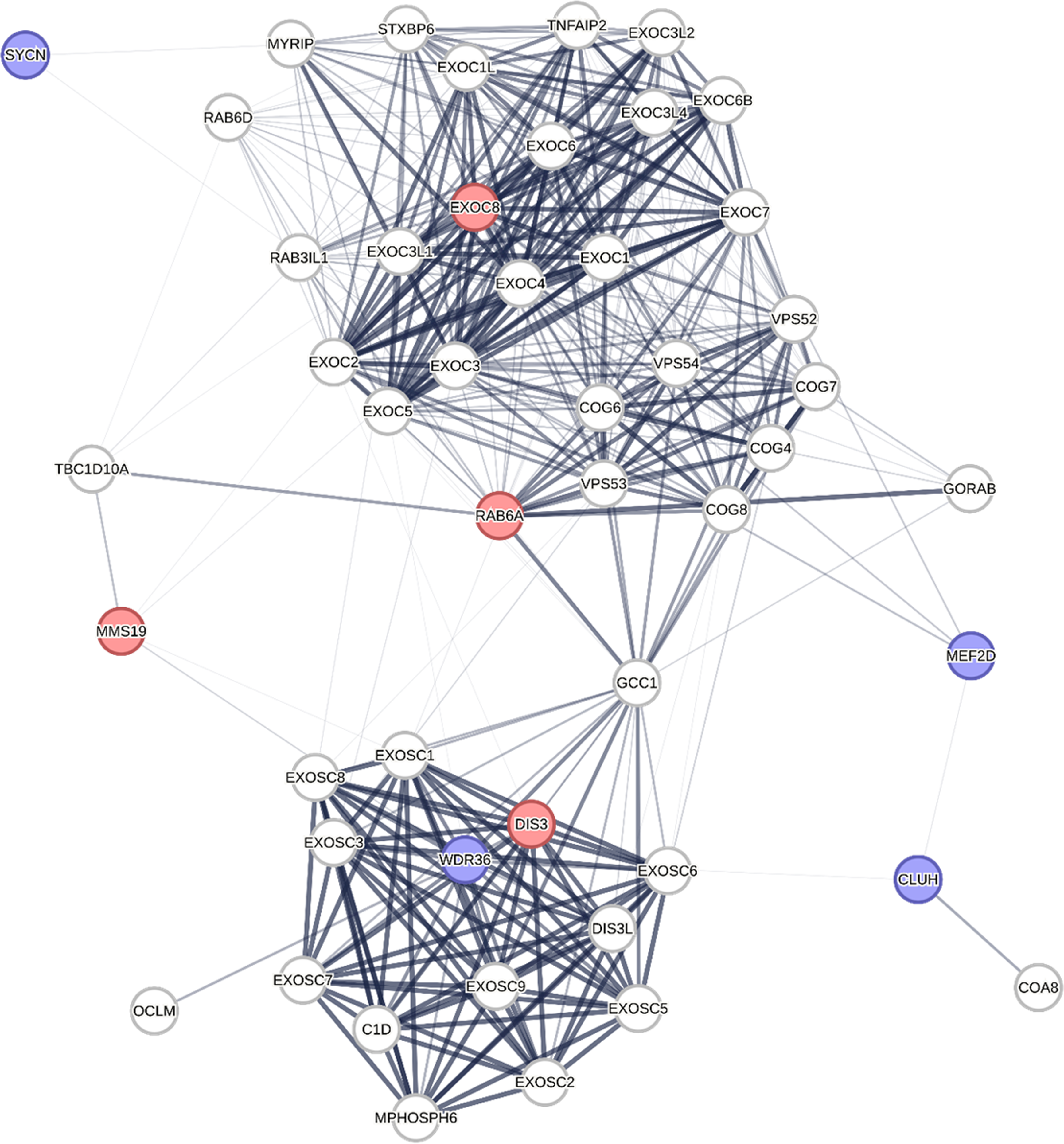
Protein-protein interaction network of up- and down-regulated proteins in the IA group compared to the control group. This network diagram represents the interactions among proteins that are differentially expressed in the IA group as compared to controls. Upregulated proteins are indicated with red nodes, including DIS3, EXOC8, RAB6A and MMS19, signifying their increased expression in IA. Downregulated proteins are shown with blue nodes, such as WDR36, SYCN, CLUH and MEF2D, representing their decreased expression. The white nodes represent proteins that are present in the network but are not differentially expressed between the IA and control groups, maintaining steady expression levels. These white nodes, while not showing changes in expression, still play essential roles within the network, potentially serving as integral components of key pathways or complexes involved in IA pathogenesis. Nodes are connected by lines indicating known or predicted protein-protein interactions, with the thickness of the lines suggesting the strength of evidence supporting the interaction. The layout of the network highlights potential key regulatory proteins that may serve as central hubs in the pathogenesis of IA.

### 3.5 Validation of LC-MS/MS Data by Western blot analysis

The Western blot analysis (Figure 5) shows differential expression of upregulated proteins between the IA group and the control group, with actin serving as a consistent loading control. Quantification of these results (Figure 6) reveals that EXOC8, RAB6A and MMS19 are significantly upregulated in the IA group compared to controls, with EXOC8 showing the highest significance (*p* < 0.01). No significant difference is observed in DIS3 expression between the groups. This validation step reinforces the reliability of the proteomic findings and the potential role of these proteins as biomarkers.

**Figure 5.**
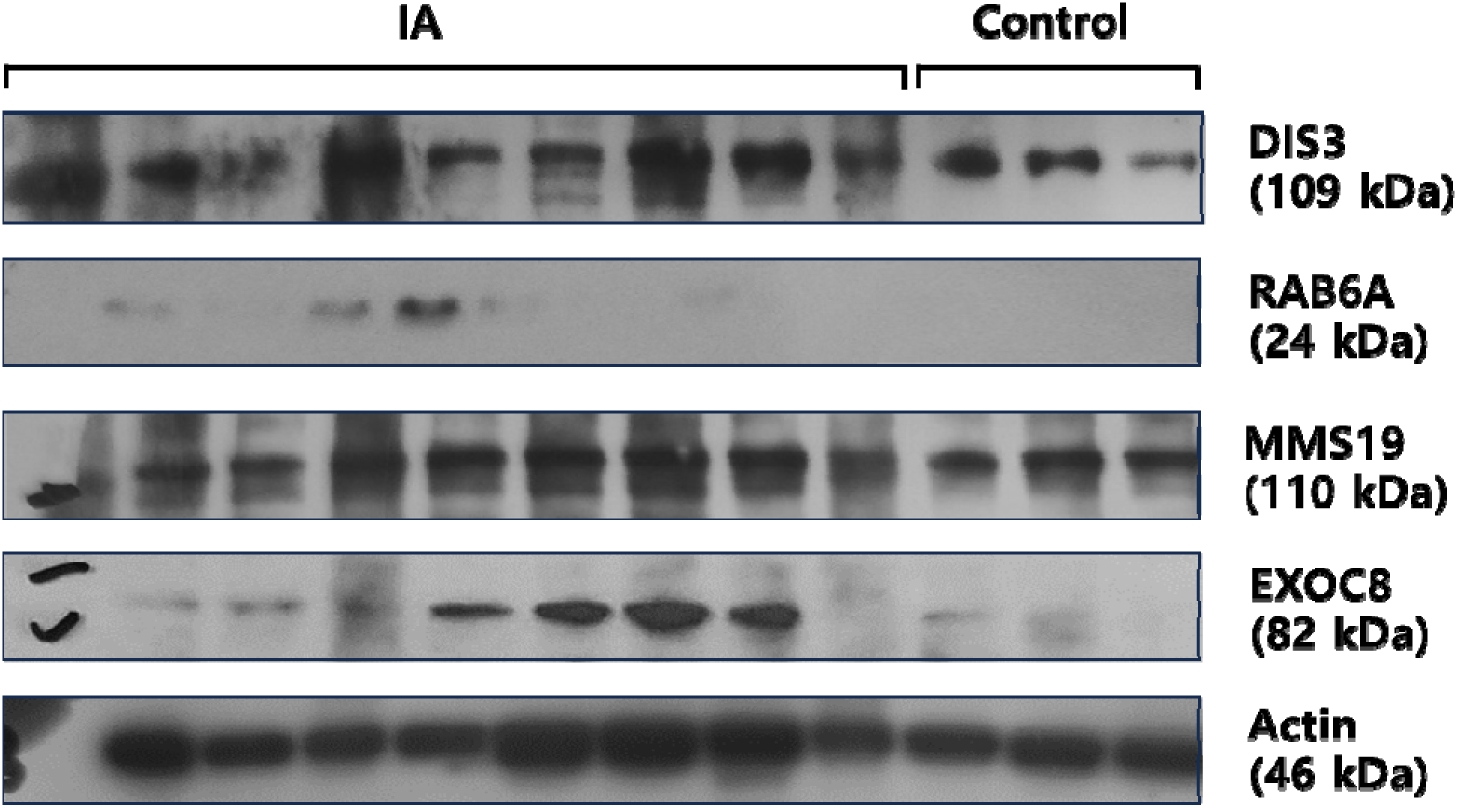
Western blot analysis displaying protein expression of candidate proteins in the study between the IA group and the control group. The presence of the target proteins is indicated by the respective bands in the lanes. The housekeeping gene, Actin (46 kDa), is used as a loading control to ensure equal protein loading across all samples and serves as an internal control for normalization purposes.

**Figure 6.**
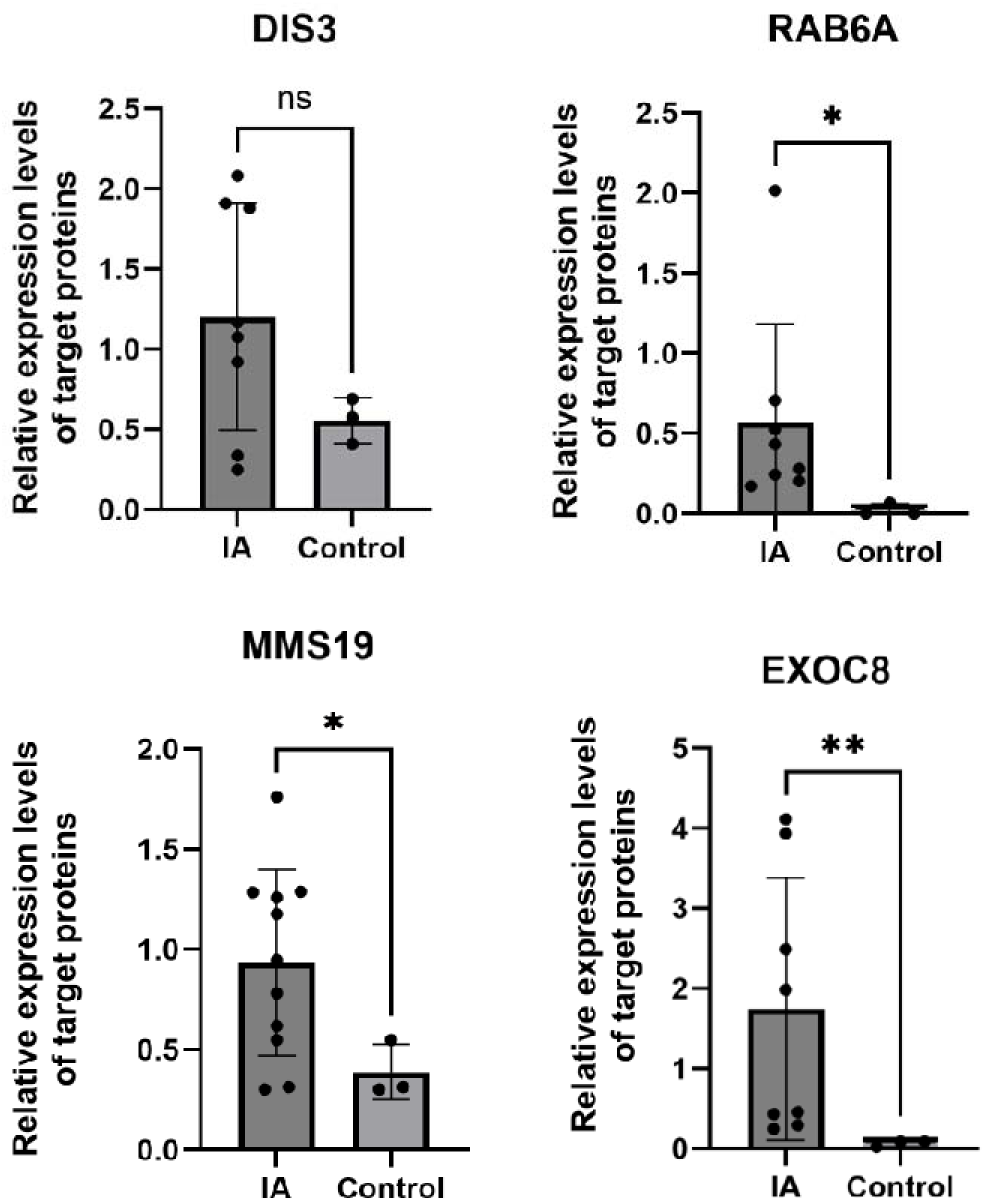
Relative expression levels of target proteins in ADPKD patients with and without intracranial aneurysms. The data are presented as mean expression levels with error bars representing the standard deviation. EXOC8 expression is significantly higher in the IA group than in the control group (*p* < 0.01). Similarly, RAB6A and MMS19 show significantly elevated expression levels in the IA group compared to the control group (*p* < 0.05 for both). No significant difference in DIS3 expression levels is observed between the two groups. Statistical significance is indicated by asterisks, with a single asterisk representing *p* < 0.05 and a double asterisk representing *p* < 0.01.

### 3.4 Correlation between candidate protein expressions and laboratory variables

Table 4 presents the correlation between candidate protein expressions and laboratory variables in the study group, highlighting significant relationships. The protein MMS19 shows a negative correlation with platelet count (r = −0.59, *p* < 0.0001) and positive correlations with total bilirubin (r = 0.397, *p* = 0.032), PT (sec) (r = 0.391, *p* = 0.047), and PT (INR) (r = 0.447, *p* = 0.021). The protein SYCN is positively correlated with platelet count (r = 0.558, *p* < 0.001) and negatively correlated with PT (sec) (r = −0.497, *p* = 0.003). MEF2D shows a positive correlation with platelet count (r = 0.432, *p* = 0.02). Finally, WDR36 is positively correlated with platelet count (r = 0.557, *p* = 0.001) and negatively correlated with total bilirubin (r = −0.368, *p* = 0.049). These correlations suggest significant associations between these proteins and specific laboratory variables in the study group.

**Table 4.**
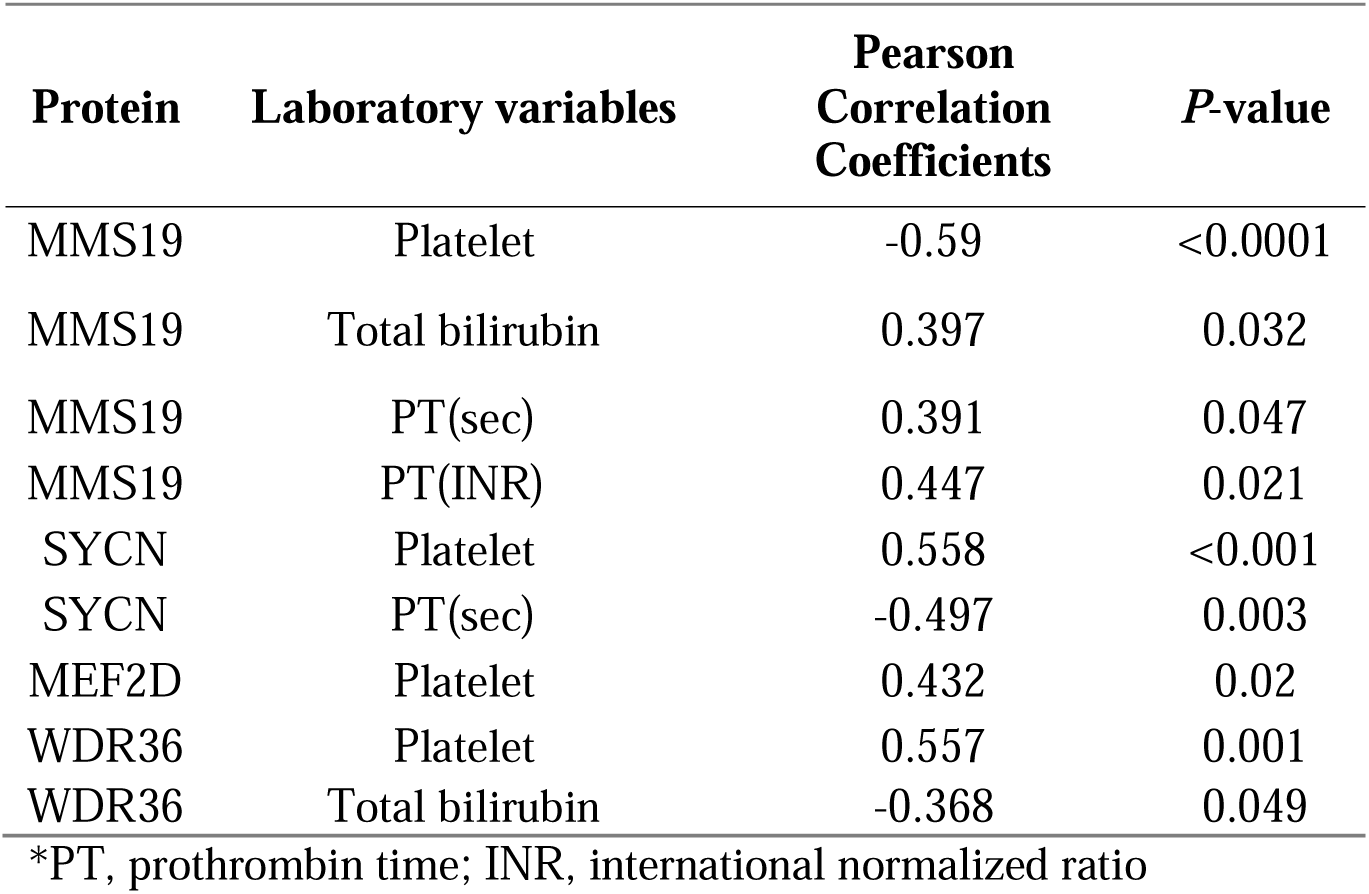
Correlation between candidate protein expressions and laboratory variables in the study group.

## 4. Discussion

IAs in ADPKD patients present a significant clinical challenge due to the increased risk of rupture, leading to subarachnoid hemorrhage, a condition with high morbidity and mortality rates. The connection between ADPKD and the development of IAs is believed to be related to abnormalities in the blood vessels and the cystic nature of the disease, which may predispose individuals to weakened arterial walls^23,24^. Molecular studies provide insight into how ADPKD contributes to vascular abnormalities. Research has shown that polycystin 1, a protein encoded by the PKD1 gene, is crucial for the structural integrity of blood vessels. Mutations in the PKD1 and PKD2 genes, which are common in ADPKD, result in altered calcium signaling in vascular smooth muscle cells (SMCs), impacting vascular reactivity and potentially leading to the development of IA^24^.

Currently, the detection and monitoring of IAs in ADPKD patients rely heavily on imaging techniques such as MRA and CTA. While these methods are effective in identifying the presence of aneurysms, they do not provide insights into the risk of aneurysm growth or rupture until potentially life-threatening symptoms emerge. This limitation underscores a critical need for novel biomarkers that can offer prognostic value, enabling early intervention and personalized management strategies for at-risk patients.

Numerous studies have explored the various causes of IAs in the general population, identifying factors such as inflammation^25,26^, tissue degeneration^27,28^, hemodynamics^29^, hormonal^30,31^ and environmental^32^. However, research specifically examining the mechanisms behind IA development in patients with ADPKD is notably limited. Polycystin-1 and polycystin-2, proteins expressed in arterial SMCs as well as in the kidneys, have been implicated in aneurysm formation due to their potential abnormalities in vascular smooth muscles^33,34^. Studies on SMC Pkd2 knockout mice reveal impaired pressure-induced constriction in cerebral arteries, linked to polycystin-2’s interaction with the actin cytoskeleton, which is vital for regulating stretch-activated currents in SMCs. This interaction is essential for maintaining pressure-induced contractility in cerebral arteries^35,36^. The higher prevalence of aneurysms in ADPKD patients may thus stem from disrupted polycystin-2 and actin interactions, resulting in compromised vascular contraction and increased susceptibility to pressure-induced vascular damage.

Despite extensive research, the search for reliable biomarkers for IAs, particularly in the context of ADPKD, has been challenging. Factors such as the heterogeneity of the disease, the complexity of aneurysm biology, and the interplay between genetic and environmental factors contribute to the difficulty in identifying specific proteins or molecular signatures that could serve as early indicators of aneurysm formation or risk of rupture.

In this study, we pioneered the identification of new proteins linked to IAs in ADPKD patients through an extensive and quantitative proteomic analysis. Utilizing advanced methods like mass spectrometry and bioinformatics, we successfully uncovered a set of proteins that had not been previously associated with this condition.

### DIS3

DIS3 is a key component of the RNA exosome complex, which is responsible for the degradation and processing of various RNA molecules^37^. Overactivity of DIS3 could lead to the aberrant degradation of RNAs^38^ that are crucial for producing proteins involved in maintaining the structural integrity of blood vessels. This disruption may weaken the vessel walls, making them more susceptible to aneurysm formation^39^. Additionally, excessive DIS3 activity might trigger cellular stress responses by accumulating defective RNAs^40^, promoting inflammation, and further compromising vascular integrity. Finally, impaired RNA processing could hinder the production of proteins necessary for vascular repair, exacerbating the risk of aneurysm development in ADPKD patients. These mechanisms highlight the potential pathogenic role of DIS3 upregulation in aneurysm formation, though further research is needed to fully understand its impact in this context.

### RAB6A

RAB6A, a small GTP/GDP-binding protein, facilitates protein transport from the endoplasmic reticulum to the Golgi apparatus and plasma membrane^41^. It plays a crucial role in macrophages’ secretion of pro-inflammatory cytokines, notably TNF-alpha^42^, which is significantly involved in the inflammatory processes weakening arterial walls and thus contributing to the development and rupture of cerebral aneurysms^43^. Shi et al. utilized microarray techniques to analyze gene expression in human IA lesions, uncovering an association between inflammation, inflammatory response, apoptosis, and IA development. They also noted an upregulation of proinflammatory genes, including TNF-α, in the walls of human IAs^44^. This connection between RAB6A and TNF-alpha underscores its critical role in the pathophysiology of IAs. Moreover, RAB6A’s role in the phenotypic modulation of SMCs under hypoxic conditions further underscores its significant impact on vascular health and aneurysm development^45^. This suggests RAB6A’s involvement extends beyond immune function, influencing vascular remodeling and structure.

### MMS19

MMS19 acts as a coactivator for estrogen receptor (ER) transcription by facilitating the incorporation of iron-sulfur into specific components of the TFIIH complex. MMS19 interacts with nuclear receptors and bind with ERα^46^. Estrogen has been found to have vasoprotective effects, including anti-inflammatory properties and the ability to promote SMC proliferation and collagen synthesis, which strengthen the arterial wall^47,48^. And this fact was clinically shown by multiple studies that show a correlation between hormon replacement therapy containing estrogen and a decreased incidence of intracranial aneurysmysmal hemmorrhages. Therefore, the activation of estrogen receptors might play a role in these vasoprotective effects. Thus, as a coactivator, MMS19 enhances the transcriptional activity of ER by interacting with it and assisting in the recruitment of the transcriptional machinery to estrogen-responsive genes, resulting in amplifying the vasoprotective effects of estrogen^49–51^.

Interestingly, our proteomic analysis revealed that MMS19 is upregulated in the IA group compared to the control group. MMS19 is known for its vasoprotective effects via activation of estrogen receptors, which initially seems paradoxical. However, we hypothesize that this upregulation may be a compensatory response to increased vascular stress and damage. The body might be attempting to utilize MMS19’s protective properties to counteract the pathological processes leading to aneurysm formation. Despite this upregulation, the complex interplay of other pro-inflammatory and pro-aneurysmal factors might be overwhelming its protective effects. This finding underscores the importance of understanding the dynamic and context-dependent nature of protein regulation and highlights the need for further research into these mechanisms.

### EXOC8

EXOC8 is one of the subunits in the exocyst complex which is made up of eight subunit, EXOC1–EXOC8. The complex has been recognized for its involvement in diverse cellular activities including exocytosis, cell growth and migration, cell polarity, cytokinesis, ciliogenesis and autophagy^52–55^. These processes are crucial for maintaining normal cellular function and structure, particularly in vascular and neural tissues. Previous research has indicated that mutations in EXOC8 play a crucial role in a neurodevelopmental disorder marked by microcephaly, seizures, and brain atrophy, which is associated with Joubert syndrome^56,57^. As a ciliary proteome component, EXOC8’s mutation is believed to contribute to Joubert syndrome by disrupting ciliary function, highlighting its importance among the many genes linked to this ciliopathy^58^. Upregulation of EXOC8 in ADPKD patients could contribute to IA formation by disrupting primary cilia function, which is crucial for maintaining vascular integrity. This disruption may impair blood vessel walls, making them more susceptible to aneurysms. Additionally, excessive EXOC8 might interfere with other cellular processes like cell migration and autophagy, further weakening the vascular structure and increasing aneurysm risk in these patients.

### Association of polycystic liver disease with IA occurrences

PCLD frequently occurs alongside PCKD in ADPKD patients, leading to a clinical situation characterized by a marked increase in liver cyst growth. When PCLD and PCKD coexist, patients often see an escalation in both the number and size of liver cysts over time, potentially impacting liver functionality^59^. Despite these developments, the liver enzyme levels and function tests for most individuals with PCLD typically remain within or near normal ranges. Nonetheless, individuals displaying symptoms often present with laboratory abnormalities, including increased levels of alkaline phosphatase, gamma-glutamyl transpeptidase, aspartate aminotransferase, and total bilirubin, with prevalence rates ranging from 15% to 70%^60^. Our analysis revealed that certain candidate proteins, such as MMS19, SYCN, MEF2D, and WDR36, exhibited correlations with laboratory test abnormalities, including platelet counts, total bilirubin levels, and PT.

As patients with ADPKD age, they typically experience an increasing frequency of symptoms and complications, indicating a progression in disease severity that parallels the intensification of disease-related manifestations^61^. A longitudinal study by R. Matsuura et al. on ADPKD patients, comparing those with and without concurrent PCLD, indicated similar platelet counts across both groups, albeit slightly lower in PCLD patients. This study also noted a steady increase in liver cyst size over time, with no reduction, and found no significant link between the volume of liver cysts and the overall liver volume^62^. This discrepancy implies that cyst growth could lead to a reduction in liver parenchyma volume, potentially affecting coagulation functions as suggested by aggravation of platelet counts and prothrombin time. Certain studies have highlighted that an increase in liver volume in PCLD patients could lead to deteriorating liver function, eventually requiring a liver transplant as the only viable treatment option^63,64^. Observational research has further established a significant association between the progression of renal cysts or renal dysfunction and the development of liver cysts^65–67^. Moreover, in the autopsy series conducted by Karhunen and Tenhu, it was found that six out of twelve cases of ADPKD with the presence of PCLD, had IAs. Conversely, none of the ten cases with isolated polycystic liver disease exhibited IAs^68^. This finding indicates that the coexistence of polycystic liver disease with ADPKD may elevate the risk of developing IAs. If a correlation exists between liver enzyme levels, affected by the volume impact of PCLD, and the incidence of IAs, assessing liver enzyme levels could serve as an indirect method to gauge the risk of IAs.

### Limitations

There are some potential limitations of our study. Firstly, the retrospective design hinders the establishment of causality between identified protein biomarkers and IA development, potentially introducing biases due to reliance on existing data. Secondly, using renal artery samples instead of aneurysmal tissue may not fully capture the specific proteomic changes at aneurysmal sites, affecting the applicability of biomarkers. Nevertheless, given that ADPKD is a systemic vascular disorder affecting multiple arterial sites, the use of renal artery samples provides a valid surrogate for studying disease-related proteomic changes. Thirdly, a larger and more diverse sample size is needed to enhance biomarker reliability. While the study confirmed increased protein expression through immunoblotting, further validation in independent cohorts is essential. Additionally, confounding factors such as treatment variations, comorbidities, and lifestyle differences were not fully addressed. Future research should incorporate prospective designs, direct aneurysmal tissue analysis, larger sample sizes, and strategies to mitigate confounding factors to validate the diagnostic potential of the identified proteins.

Our study identified several differentially expressed proteins in the renal arteries of ADPKD patients, distinguishing those with IAs from those without. Key proteins, including DIS3, RAB6A, MMS19, and EXOC8, were upregulated in patients with IAs, while CLUH, SYCN, MEF2D, and WDR36 were downregulated. In conclusion, this study enhances our understanding of the proteomic changes associated with IAs in ADPKD patients, identifying potential biomarkers for early diagnosis. Furthermore, the presence of PCLD in individuals with ADPKD may act as a marker for disease advancement. This progression could also imply an increased risk of aneurysm formation, as both previous research and the current study have associated the coexistence of PCLD and ADPKD with a higher incidence of IA. Consequently, regularly assessing liver enzyme levels in these patients might offer an indirect approach to estimating their likelihood of developing IA. Further investigation and interdisciplinary efforts are needed to translate these findings into clinical practice, improving patient outcomes and advancing personalized medicine for ADPKD-related IAs.

## Data Availability

The datasets generated and analyzed during the current study are available from the corresponding author on reasonable request. Due to privacy restrictions, some data may be subject to ethical or legal considerations, and access may be granted upon approval of the appropriate institutional review board.

http://www.ebi.ac.uk/pride/archive/projects/PXD043129

## Non-standard Abbreviations and Acronyms

ADPKD: Autosomal Dominant Polycystic Kidney Disease
AFA: Adaptive Focused Acoustics
BMI: Body Mass Index
CVA: Cerebrovascular Accident
CTA: Computed Tomography Angiography
DM: Diabetes Mellitus
eGFR: Estimated Glomerular Filtration Rate
EXOC8: Exocyst Complex Component 8
FDR: False Discovery Rate
HD: Hemodialysis
IA: Intracranial Aneurysm
KT: Kidney Transplantation
LC-MS/MS: Liquid Chromatography-Tandem Mass Spectrometry
MRA: Magnetic Resonance Angiography
PCKD: Polycystic Kidney Disease
PCLD: Polycystic Liver Disease
PVDF: Polyvinylidene Difluoride
SMC: Smooth Muscle Cell

## Acknowledgement

None

## Supplemental Material

Supplementary Methods 1

Supplementary Figure S1

