## Supplementary Figure S1 for "Exploring the Molecular Pathways of Intracranial Aneurysm Formation in Autosomal Dominant Polycystic Kidney Disease Using Proteomic Analysis"

**Supplementary data**

**Supplementary Figure S1.**

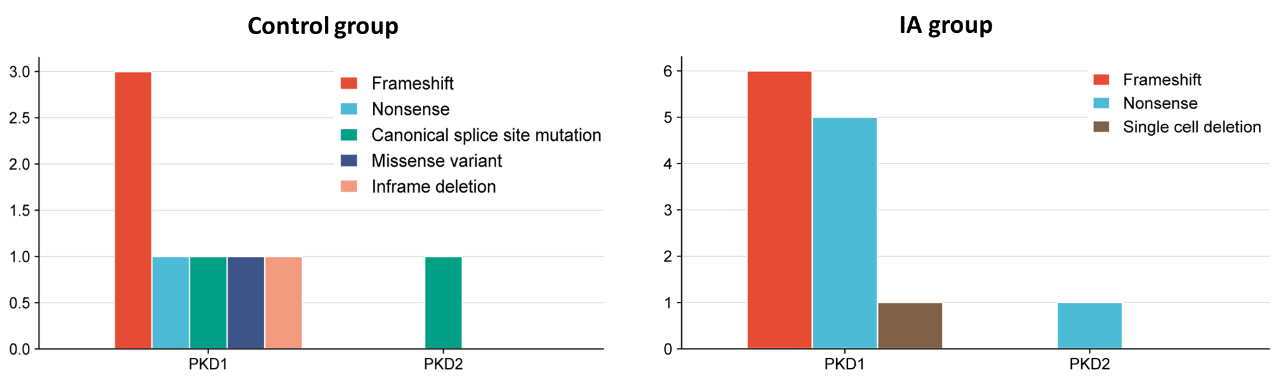

**Figure S1.** Frequency of mutation types in PKD1 and PKD2 genes across the groups. The bar graphs illustrate the distribution of different mutation types within each group. In the control group, frameshift and nonsense mutations are predominant in the PKD1 gene, with a single cell deletion also present. PKD2 mutations in the control group are all nonsense mutations. In the IA group, a diverse range of mutations is observed in the PKD1 gene, with the PKD2 gene showing only canonical splice site mutations.
