## Supplementary Methods 1 for "Exploring the Molecular Pathways of Intracranial Aneurysm Formation in Autosomal Dominant Polycystic Kidney Disease Using Proteomic Analysis"

**Supplementary data**

**Supplementary Methods 1.** Description of the method used in whole exome sequencing

WES was performed following the CAP/CLIA validated standard operating protocol. Briefly, exome capture was performed using xGen Exome Research Panel v2 (Integrated DNA Technologies, Coralville, Iowa, USA) and sequencing was performed using the NovaSeqX platform (Illumina, San Diego, CA, USA) as 150bp paired-end reads. Sequencing data were aligned to the GRCh38 human reference genome using BWA-MEM and processed for variant calling by GATK v4.2.14. Variants were then annotated by Ensembl Variant Effect Predictor (VEP) and filtered and classified by EVIDENCE v4 following the American College of Medical Genetics and Genomics (ACMG) guideline. The filtered and classified variant list was manually reviewed by medical geneticists and physicians. The most likely variants that can explain the patient’s phenotype were selected for reporting.
